# Usability and Accuracy of the SWIFT-ActiveScreener: Preliminary evaluation for use in clinical research

**DOI:** 10.1101/2023.08.24.23294573

**Authors:** Jenny J.W. Liu, Natalie Ein, Julia Gervasio, Bethany Easterbrook, Maede S. Nouri, Anthony Nazarov, J. Don Richardson

**Affiliations:** Western University Schulich School of Medicine & Dentistry; Lawson Health Research Institute; McMaster University Department of Psychiatry and Behavioural Neurosciences

## Abstract

Systematic reviews (SRs) employ standardized methodological processes for synthesizing empirical evidence to answer specific research questions. These processes include rigorous screening phases to determine eligibility of articles against strict inclusion and exclusion criteria. Despite these processes, SRs are a significant undertaking, and this type of research often necessitates extensive human resource requirements, especially when the scope of the review is large. Given the substantial resources and time commitment required, we investigated a way in which the screening process might be accelerated while maintaining high fidelity and adherence to SR processes. More recently, researchers have increasingly turned to artificial intelligence-based (AI) software to expedite the screening process. This paper evaluated the accuracy and usabiity of a novel, machine learning program, Sciome SWIFT-ActiveScreener (ActiveScreener) in a large SR of mental health outcomes following treatment for PTSD. ActiveScreener exceeded the expected 95% accuracy of the program to predict inclusion or exclusion of relevant articles, and was reported to be user friendly by both novice and seasoned screeners. Our results showed that ActiveScreener, when used appropriately, may save considerable time and human resources when performing SR.

## Introduction

Systematic reviews (SRs) are the current standard to collate and synthesize empirical evidence and evaluate trends across a specific body of literature in response to specific research questions. SRs involve strict structured and formal methodological processes (Gough et al., 2020; White et al., 2012). Standardized protocols, such as the Preferred Reporting Items for Systematic Reviews and Meta-Analysis (PRISMA) offer researchers a roadmap to conducting SRs with rigour and fidelity (Page et al., 2021). In addition, formal guides established by Cochrane provide additional evaluation criteria in order to provide appropriate context for the interpretation of study data in various research settings (Higgins et al., 2021). Despite these protocols, SRs continue to be a huge undertaking due to extensive resource requirements. Depending on the scope of review and precision of search terms used, researchers may review tens of thousands of articles during various stages of screening. Therefore, given the substantial resource and time commitment required to complete the screening phases for SRs it is crucial to investigate opportunities which may accelerate the screening process. In this paper, we evaluated ActiveScreener in terms of its accuracy and usability in a large SR of mental health outcomes following treatment for PTSD. ActiveScreener was selected namely for its departure from programs that uses AI to identify records, and instead, uses machine learning to build a predictive algorithm to reduce time spent in screening phases of SRs.

The screening phases of a SR includes de-duplicating search outputs across multiple database, and screening title and abstracts and full text (Page et al., 2021). During these steps, researchers examine each article against strict inclusion and exclusion criteria in order to determine its eligibility for inclusion in the SR. To ensure standards of quality, more than one individual must screen the same article independently at each screening stage, with the reliability between screeners calculated and reported as part of the SR (Belur et al., 2021). Altogether, screening phases can take hundreds of hours for each individual reviewer involved.

Artificial intelligence-based (AI) softwares such as COVIDENCE (Veritas Health Innovation, n.d.), CUREDATIS (Research Solutions, 2023), and Sciome SWIFT-ActiveScreener (Howard et al., 2020) have been developed help expedite SR screening. For the purpose of this paper, AI programs refer to programs that are enabled to perform tasks that normally require human intelligence during in the context of conducting a SR. While they do not eliminate human involvement in the screening process, each program may reduce time and resources spent using various proprietary solutions. For example, COVIDENCE aids clinical research reviews with its ability to distinguish between articles which are randomized controlled trials (RCTs) versus non-RCTs.

ActiveScreener is a novel, machine learning and web-based AI software for SRs. ActiveScreener uses a pretrained machine learning program to identify and prioritize articles for screening (Howard et al., 2020). Based on user feedback via patterns of screening, ActiveScreener uses its pretrained algorithm to build a model estimating articles for inclusions versus exclusions, trimming screening time and effort by nearly 70% (Howard et al., 2020). Past research utilizing ActiveScreener have found the algorithm to work well in reviews involving the physical health literature (Elmore et al., 2020; Lam et al., 2019).

### Aims

Despite indications of past use in health reviews, there is little evidence for how ActiveScreener may perform in evaluations of mental health and treatment outcomes. Further, the precision of the estimation model remains unclear. In this paper, we set out to evaluate the precision and usability of ActiveScreener in conducting screening for a mental health treatment SR (Liu et al., 2021). Specifically, in Part 1, we formally evaluated the accuracy of its predictive model relative to the actual outcomes of screening conducted by individual human screeners, and in Part 2, we collected informal feedback regarding the usability of ActiveScreener amongst a cohort of screeners.

## Part 1 – Accuracy

## Methods

### Procedure

Eighteen screeners were trained to identify articles for inclusion and exclusion and on the use of ActiveScreener for a meta-analysis and systematic review (for more details on this project see Liu et al., 2021). A total of 10002 references required review at the title and abstract stage. ActiveScreener accuracy statistics were set at 95% resulting in 5390 of these references to be reviewed by screeners. Once screening reached 95% accuracy, all screeners stopped. At this stage, data consisting of the screening results for the 5390 references reviewed by screeners and the remaining 4612 references reviewed by the ActiveScreener AI were exported. Accuracy statistics were then reset to 100% prompting the screeners to continue screening the remining 4612 references. Data was once again exported. Screening results for the 4612 references from ActiveScreener and the screeners were then compared.

### Data Analysis

A confusion matrix and statistics were generated and used to evaluate the predictive accuracy of ActiveScreener across three classes. The three classes were Included (represents references identified as meeting inclusion criteria), Excluded (representing references identified as meeting exclusion criteria), and Conflicted (representing disagreement on whether the reference should be included or excluded). Analyses were performed in R-Studio, using the tidyverse (Wickham et al., 2023), stringr (Wickham, 2022), and caret (Kuhn, 2008) packages. Results are reported for only the title and abstract screening stage.

## Results

The multiclass confusion matrix for 4612 references is presented in Table 1. As shown, both the screeners and the ActiveScreener AI accurately identified 1365 included references, 2528 excluded references, and 622 conflicted references. For 97 references, the screeners identified these references as included, while the ActiveScreener AI identified these references as conflicted.

**Table 1.** Confusion Matrix (n = 4612)

|  | Actual |  |  |  |
| --- | --- | --- | --- | --- |
| Predicted |  | Conflicted | Excluded | Included |
|  | Conflicted | 622 | 0 | 97 |
|  | Excluded | 0 | 2528 | 0 |
|  | Included | 0 | 0 | 1365 |

Notes. Actual = screeners; Predicted = ActiveScreener

Overall, accuracy was 97.9%, 95% CI [0.97, 0.98], *p* < .001. Interrater reliability was reported with Kappa [Fleiss and Conger; 0.96]). Sensitivity for the three classes were: Included (0.93), Excluded (1.00), and Conflicted (1.00). Specificity for the three classes were: Included (1.00), Excluded (1.00), and Conflicted (0.98).

## Part 2 - Usability

## Methods

### Respondents

Eighteen screeners completed a survey around the usability of ActiveScreener. All respondents were paid employees or unpaid volunteers of the MacDonald Franklin Operational Stress Injury Research Centre (MFOSIRC).

### Measures

#### Demographic

Demographic information included: (1) the respondents’ role within MFOSIRC, (2) whether the respondents conducted or assisted on a systematic review or meta-analysis prior to their placement at the MFOSIRC, (3) respondents’ level of experience with systematic reviews or meta-analyses (e.g., intermediate), and (4) types of software used by respondents for screening for systematic reviews or meta-analyses.

#### ActiveScreener User Experience Survey

This survey was created by authors (J.J.W.L & A.N) to capture respondents experiences using ActiveScreener. The survey consisted of 12 items (statements or questions) related to usability of ActiveScreener for screening (e.g., “SWIFT Active Screener is easy to use”, “SWIFT Active Screener software was easy to learn”). Nine statements were quantitative, and three questions were qualitative. Of the quantitative items, eight statements were rated on a 5-point Likert scale, ranging from strongly agree to strongly disagree, and one question was rated on a 5-point Likert scale, ranging from very confident to not at all confident. The qualitative items included three open-ended questions capturing information related to features of ActiveScreener the respondents enjoyed, any challenges experienced while using ActiveScreener, and any suggestions the respondents had to improve ActiveScreener.

### Procedure

All respondents received an email with the link directing them to the online survey. Respondents were asked to complete both the demographic information and the ActiveScreener User Experiences Survey online via Google Forms. Data was collected in April 2022. No direct compensation was given for participating in this study. However, many of the respondents were paid employees of the MFOSIRC and completed the survey during working hours thereby receiving nominal monetary compensation for the time spent participating. For unpaid volunteers, the time spent completing this survey was included in their volunteer hours for which they are provided a letter of recognition.

### Data Analysis Plan

Both quantitative and qualitative data was used to provide descriptive information related to the respondents’ experiences using ActiveScreener. For qualitative data, common themes were extracted from responses provided regarding enjoyable features of the software, challenges with ActiveScreener and suggested improvements.

## Results

### Quantitative Data

All 18 respondents completed all nine quantitative items. All respondents (100%) either agreed or strongly agreed that: their training needs were met; ActiveScreener was easy to learn; they felt confident using ActiveScreener; and they would recommend ActiveScreener for use in other reviews. Nearly all respondents reported either agreeing or strongly agreeing that: ActiveScreener was easy to use (94.4%); and ActiveScreener had a user-friendly interface (94.5%). The majority of respondents (88.9%) also reported that they either agreed or strongly agreed that ActiveScreener had all the features needed for adequate screening. Of the eight respondents who had prior experience with other screening programs or tools, seven respondents (87.5%) rated that they either agreed or strongly agreed that they preferred ActiveScreener over other programs. With regards to the experience of technical or system-related glitches, respondents varied in their perspectives, with 44.5% of respondents indicating that they experienced no technical or system-related glitches (either agreed or strongly agreed), while 22.2% indicated experiencing technical or system-related glitches (disagreed). Results for each survey items are reported in Table 2.

**Table 2.**
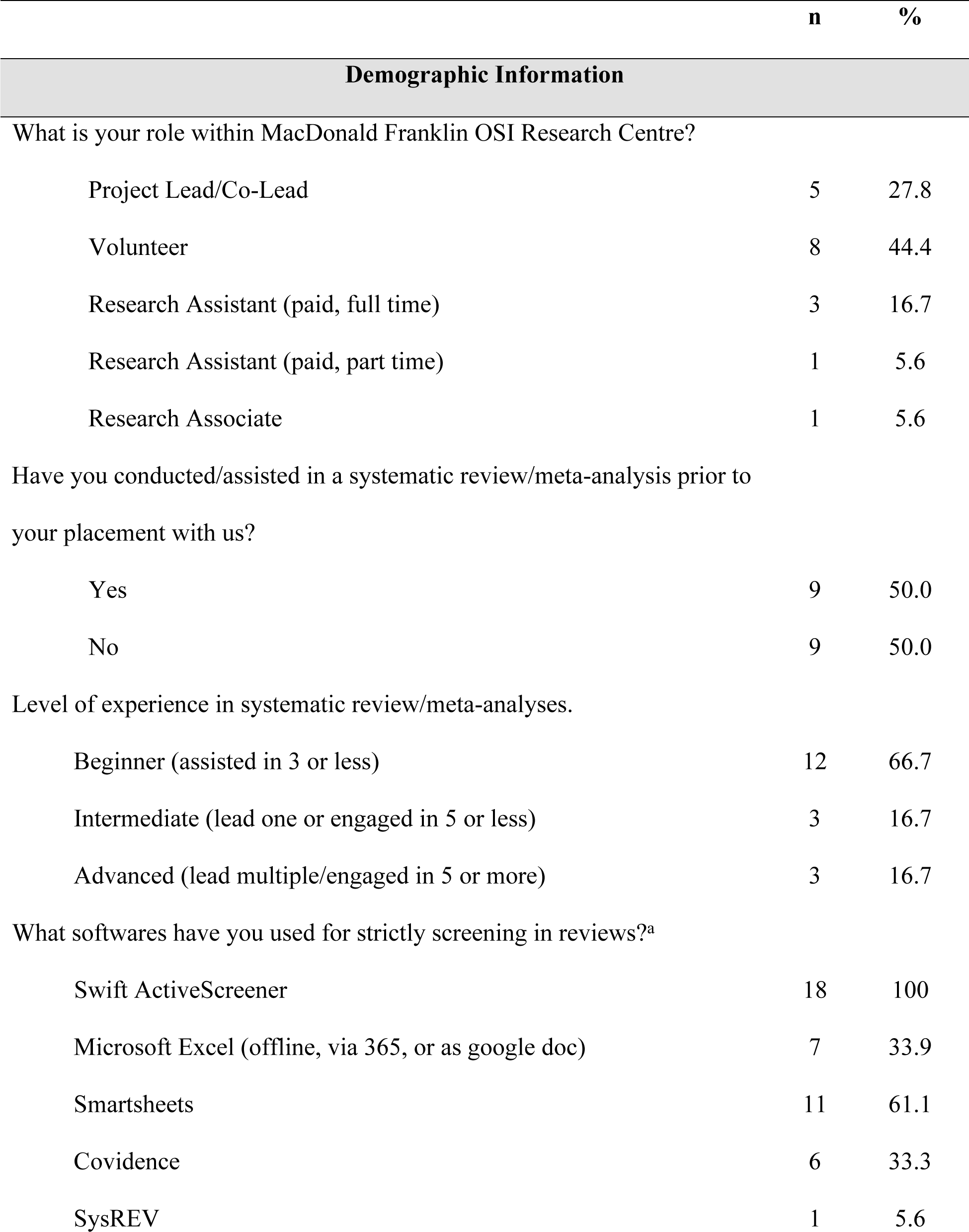
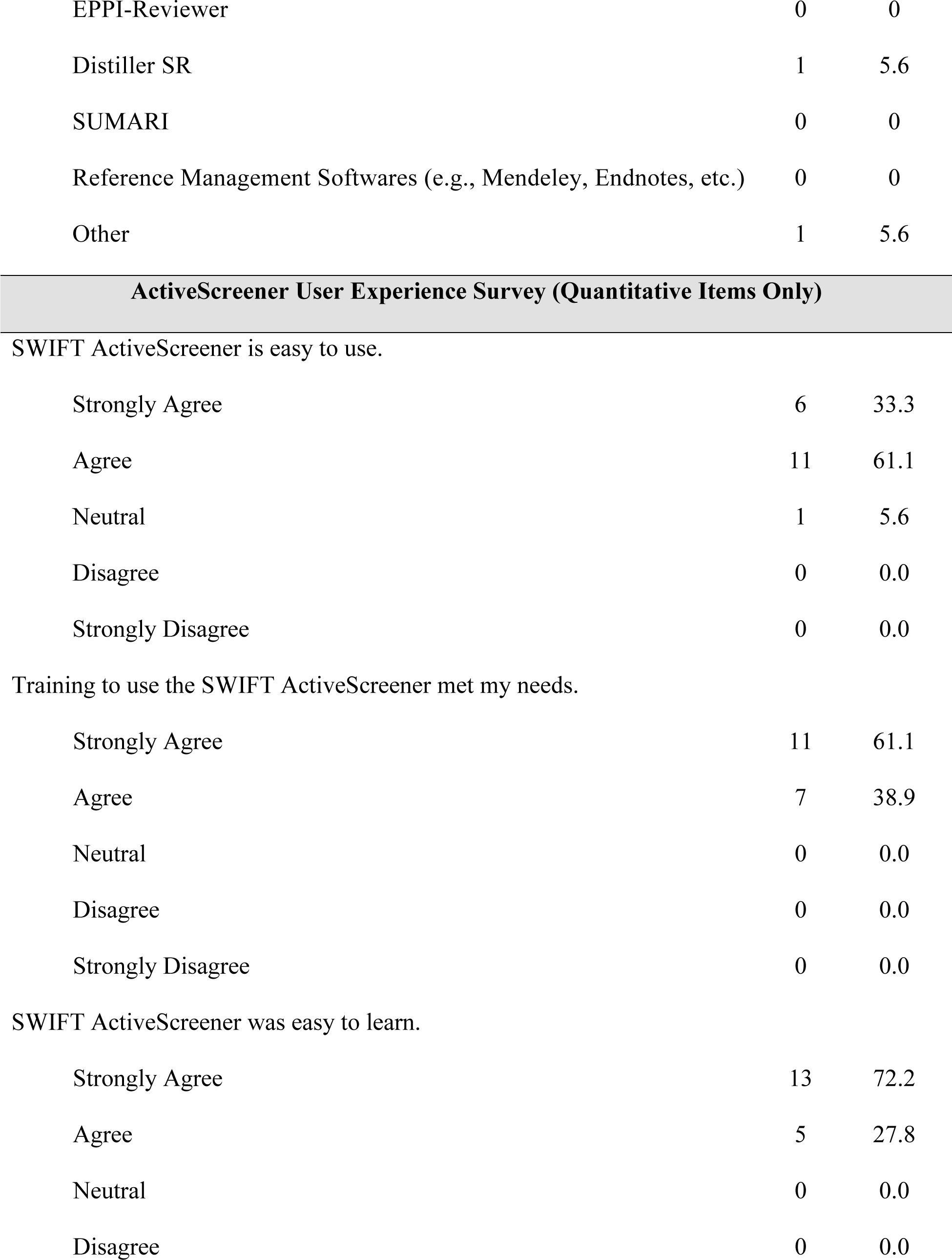
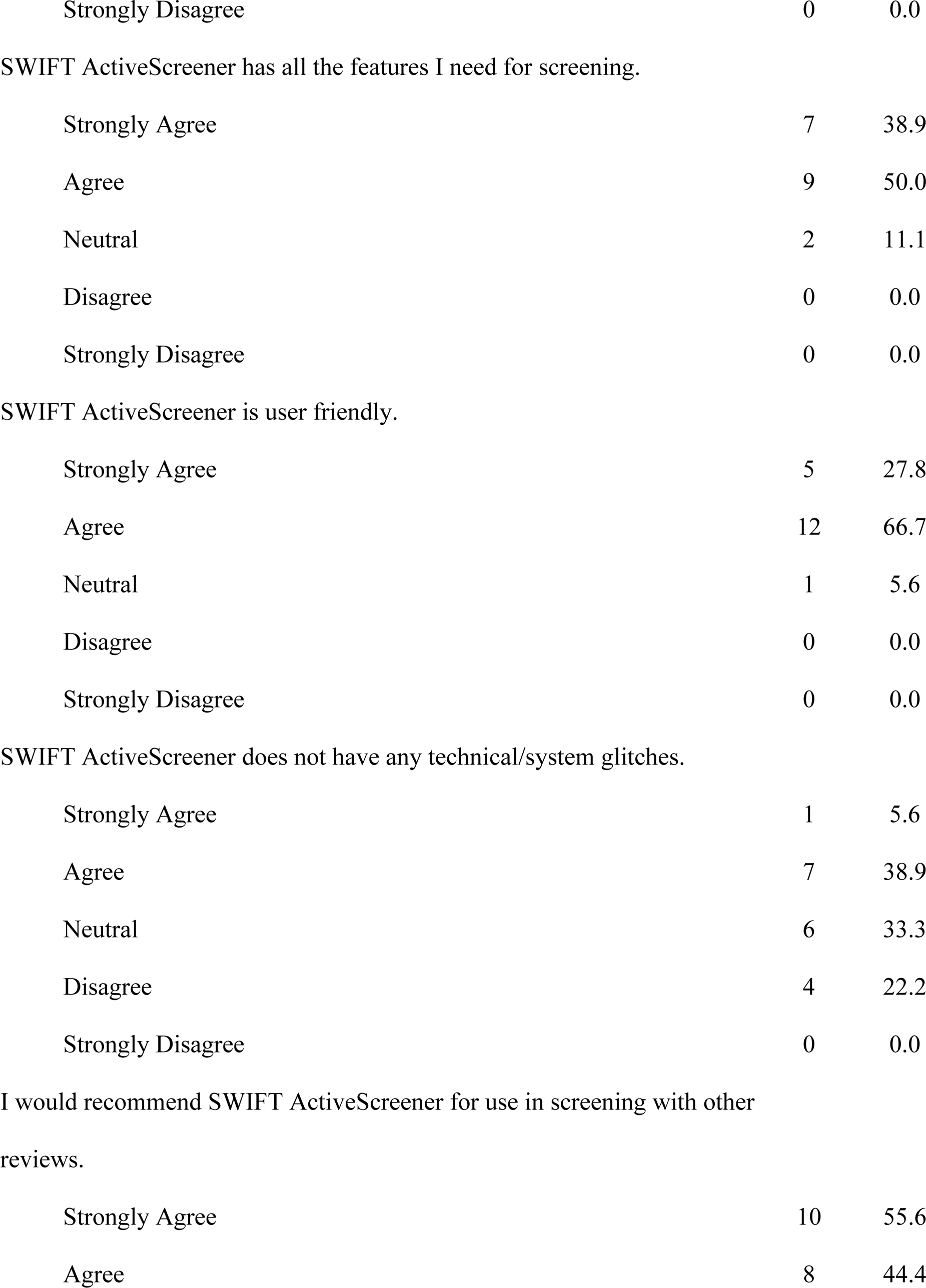
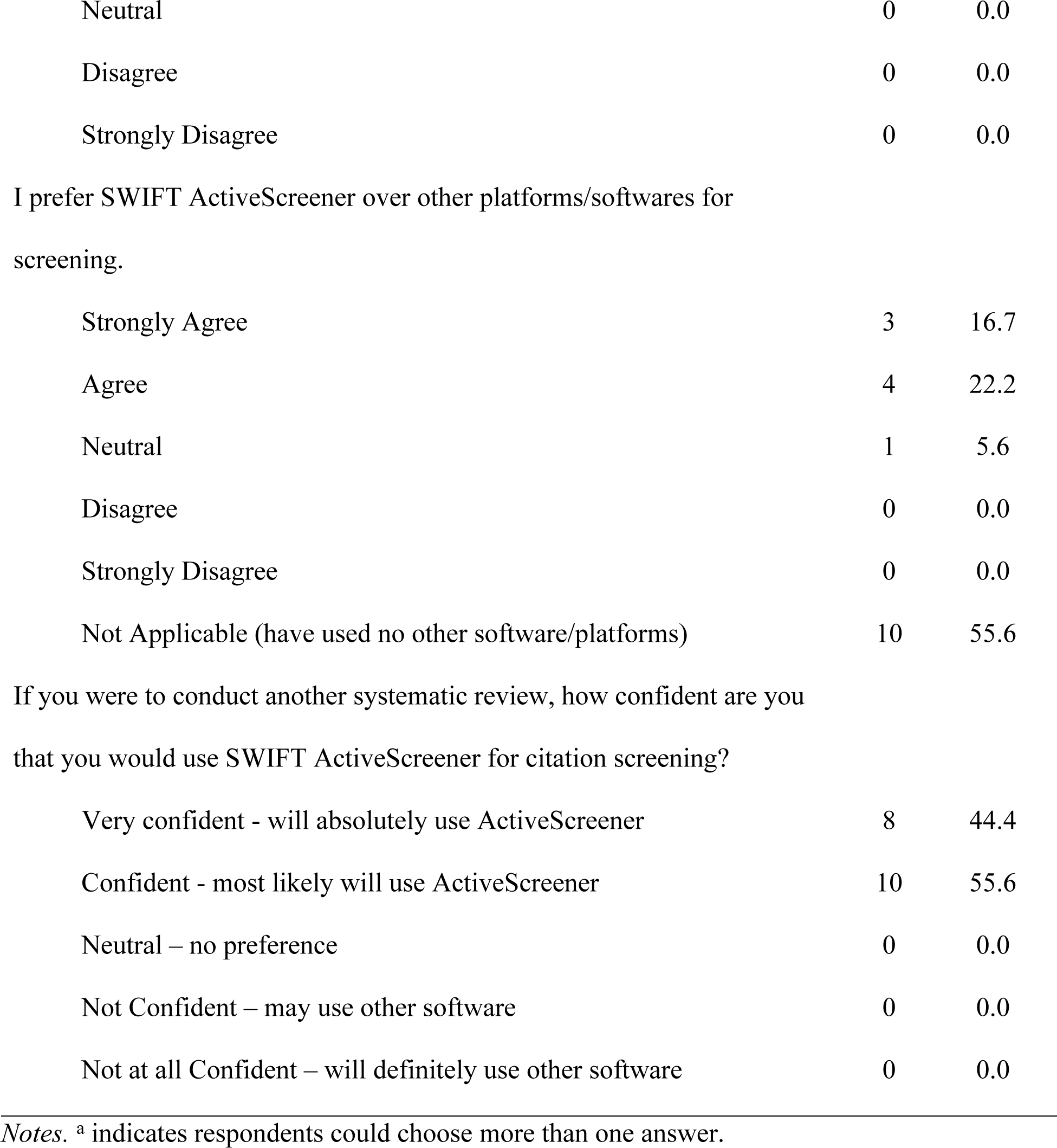
Respondent Data Across Measures (N = 18)

### Qualitative Data

#### Features Enjoyed

For the question capturing the features of ActiveScreener enjoyed most by respondents, three primary themes emerged from the data (see Table 3 for quotes).

**Table 3.**
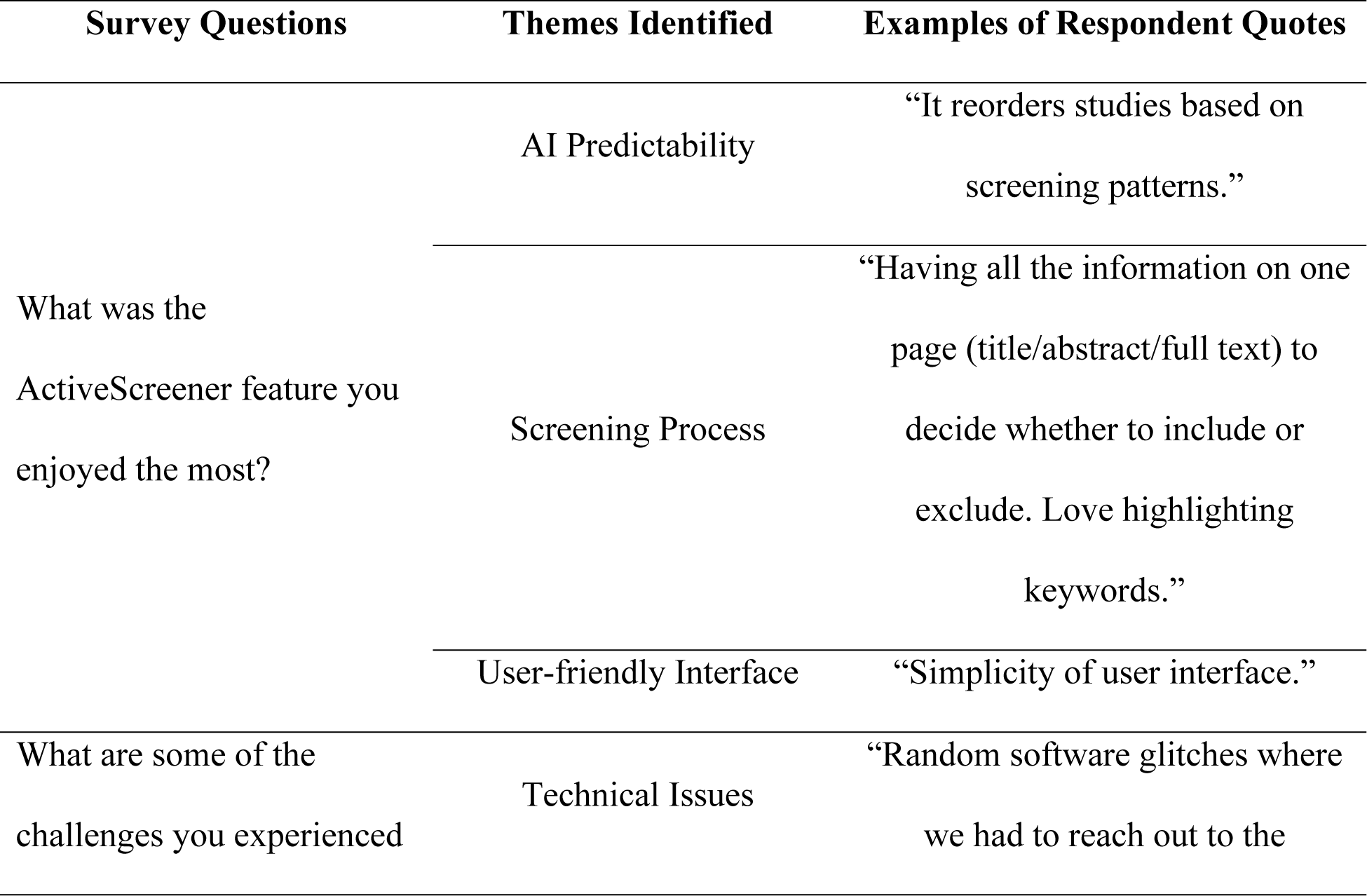
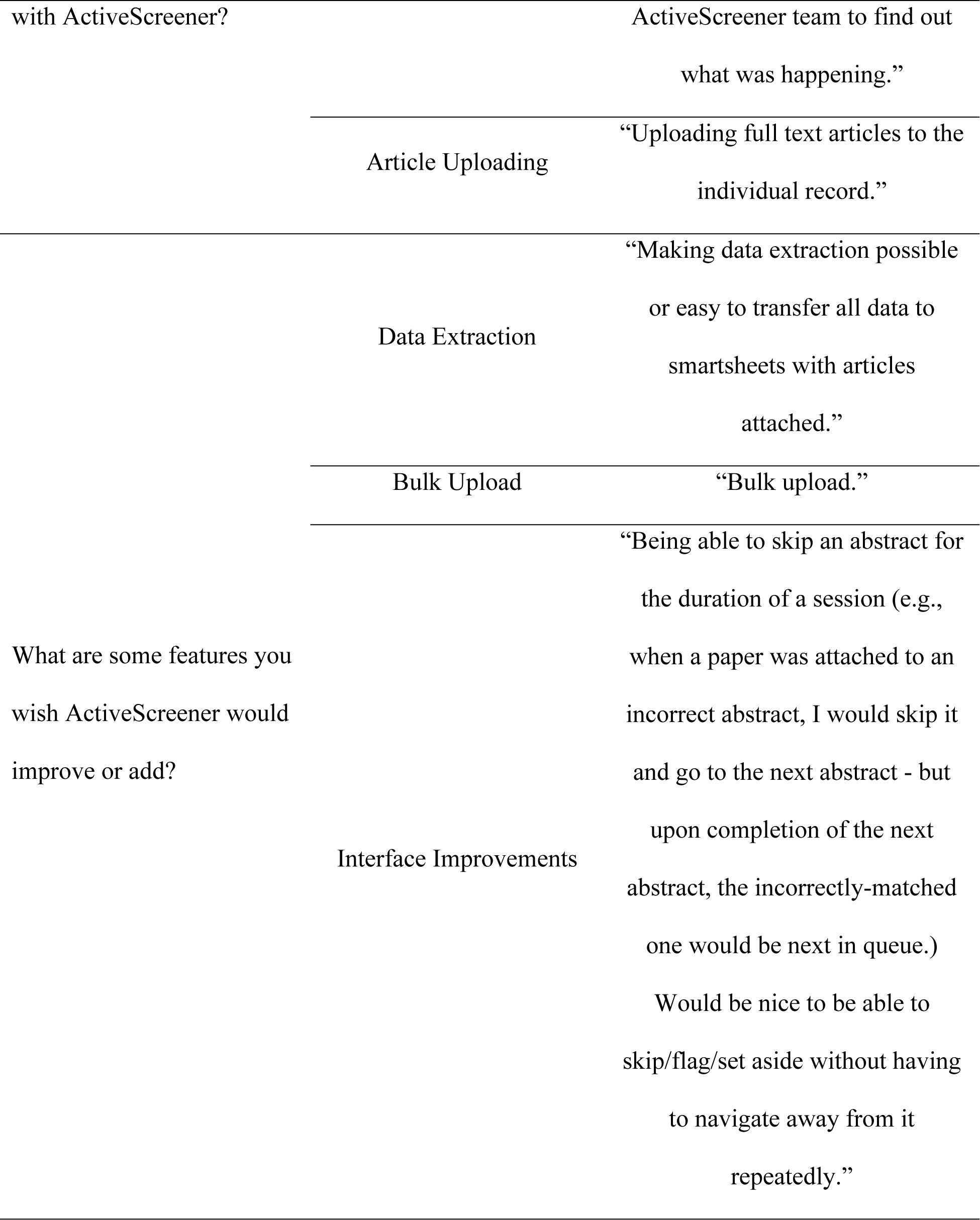
ActiveScreener User Experience Survey Qualitative Feedback.

##### AI Predictability

Respondents noted that ActiveScreener accelerates the screening process through predictive capabilities. Specifically, ActiveScreener reorders references based on individual patterns of inclusion and exclusion such that likely included articles are pushed to the top of the screening list.

##### Screening Process

Respondents noted that ActiveScreener makes the screening process easier and faster. Specifically, all the information required for screening is available on one page including the article title, abstract, full text, and inclusion and exclusion criteria. This allows the screener to evaluate the article quickly.

##### User-friendly Interface

Respondents noted that ActiveScreener has a user-friendly interface. For example, respondents noted ease of use and ability to access ActiveScreener from any device as a positive feature of this software.

#### Challenges

For the question capturing any challenges experienced by respondents, two primary themes emerged from the data (see Table 3 for quotes).

##### Technical Issues

Respondents noted that they encountered some technical difficulties and glitches while using ActiveScreener. For example, connection loss specific to the ActiveScreener website or processing or loading speeds were commonly described.

##### Article Uploading

Respondents noted that uploading articles individually to each reference is time consuming and could result in errors such as a mismatch of articles to references.

#### Suggested Improvements

For the question capturing suggested improvements or additions to the program, three primary themes emerged from the data (see Table 3 for quotes).

##### Data Extraction

Respondents noted that they would have liked the ability to either extract data directly within ActiveScreener or be able to export the included references with attached articles to other formats (e.g., SmartSheets).

##### Bulk Upload

Respondents noted that they would like the ability to upload articles to references in bulk as opposed to one at a time.

##### Interface Improvements

Respondents noted potential improvements to the user interface. For example, navigation opportunities, keeping a session counter of screened articles, and ability to flag references with incorrect articles attached.

## Discussion

In our study, we found that ActiveScreener performed above its expected 95% accuracy in prediction and was found to be user friendly by both novice and seasoned screeners. Consistent with past evidence that the effectiveness of this program can reduce screening time and effort by nearly 50% (Howard et al., 2016), we observed similar results with a large-scale review of PTSD treatment outcomes.

Regarding its accuracy, our confusion matrix results indicated that when testing against a large-scale SR which included over 10000 articles screened in the title and abstract phase, ActiveScreener performed better than expected in its predictive algorithm. While the software was expected to reach 95% accuracy, the actual accuracy of its machine learning model in our review exceeded 95% (97.9%). Further, of the categories of accuracy examined, discrepancies between the predictive algorithm and actual human screening outcomes were minimal. Specifically, there were no discrepancies between human screeners and the ActiveScreener AI with respect to articles that should be excluded from the SR. Only a small number of discrepancies were found between human screeners that indicated articles should be included while the ActiveScreener AI predicted that the articles would be conflicted (i.e., predicted multiple human screeners would disagree on inclusion and exclusion) based on prior trends in human screening. This means that no studies that the ActiveScreener AI predicted to be included resulted in exclusions by screeners. Thus, these accuracy statistics indicate that ActiveScreener is a reliable and rigorous platform to accelerate screening at the title and abstract phase of SRs, especially when utilizing its predictive algorithm function.

In examining user feedback amongst a group of screeners, we found that ActiveScreener was endorsed as easy to learn and easy to use. However, user feedback also noted that there were software glitches, such as the platform being unavailable from time to time, as well as glitches when uploading articles and using other features. While these challenges do not undermine its use, they provide areas of opportunity for ActiveScreener programmers to consider for future research and development. Further, to reduce human resources during screening, ActiveScreener should consider implementing new features such as bulk upload and templates for subsequent data extraction directly within the platform. Both would reduce the need for switching between programs when conducting reviews, and thereby reduce human resource requirements as well as potentials for errors.

## Conclusion

In considering the merits of ActiveScreener, it should be noted that the software’s machine learning algorithm is reliant on the rigour of training and the strength of screeners that it bases its user feedback on. As such, users must conduct training and screening with care. In particular, the clarity in which inclusion and exclusion criteria may be applied during the initial screening stages is of vital importance in building the accuracy of the predictive model. Thus, researchers are encouraged to spend considerable time to ensure the inclusion and exclusion criteria are clearly understood and reliably applied by all screeners during the project training stages. In addition, another time-saving feature of ActiveScreener, the deduplication function for uploading references can benefit from further development as it currently limits the deduplication to texts only, and does not extend to cover punctuation. Depending on the database, references may be exported with variable punctuations, which is not covered by the feature, resulting in many duplicate references when screening. However, it should be noted that this can easily be solved with work-arounds, such as manually combining search yields on *r* with generated codes that deduplicates references prior to uploading on ActiveScreener. Finally, it is important to note that ActiveScreener’s program to accelerate the screening stage is only currently relevant at the title/abstract stage and excludes further reviews of full-text. And thus, current study findings and the potential time and resource savings are only applicable to the initial screening phase of SRs. Taken together, ActiveScreener appears to be a user friendly and accurate platform for SRs, and when used appropriately, may save considerable time and human resources during the initial screening process.

## Data Availability

The data underlying the results presented in the study are available by request.

## Notes

### Competing Interest Statement

The authors have declared no competing interest.

### Clinical Protocols

https://www.crd.york.ac.uk/prospero/display_record.php?ID=CRD42021245754

### Funding Statement

This work is supported in part by the Atlas Institute for Veterans and Families

### Author Declarations

This work is part of meta-analysis which is exempt from research ethics.

